# Excess Mortality Resulting from COVID-19 in Turkey during 2020-2021: Regional and Time-Based Analysis

**DOI:** 10.1101/2023.05.04.23289498

**Authors:** Salih Keskin, Gül Ergör

## Abstract

Turkey experienced substantial excess mortality in 2020 and 2021 related to the COVID-19 pandemic. Methods used to estimate excess mortality vary, making comparisons difficult. This study assessed the impact of the COVID-19 pandemic in Turkey, using the TURKSTAT data which became available on February 23, 2023. We applied a quasi-Poisson model to estimate excess mortality during 2020–2021, comparing excess mortality by time periods and socioeconomic factors. During 2020–2021, Turkey experienced 72,886 excess deaths in 2020 (P-score 16.8%) and 125,540 in 2021 (P-score 28.5%). An additional 80 excess deaths per 100,000 people were recorded in 2020 and 143 in 2021. Excess all-cause mortality varied across socioeconomic levels, with notable social disparities in pandemic deaths as the highest rates were observed in the lowest socioeconomic group. This study highlights the importance of a comprehensive approach to address the diverse impacts of the pandemic on health and well-being while considering socioeconomic disparities, and potential areas for improvement in data collection and reporting.

## Introduction

The unprecedented impact of COVID-19 on global mortality rates has been widely recognized, with Turkey being no exception. Effective risk communication and the dissemination of accurate information are crucial in managing disasters such as pandemics. However, limited data availability and a lack of transparency in data sharing have hindered the accurate assessment of COVID-19’s impact on mortality rates in Turkey. The government’s restrictive data policy has raised concerns among researchers and health experts, delaying the release of Turkey’s 2020 and 2021 death data until February 23, 2023, and impeding their ability to track the pandemic’s effects (1,2). Furthermore, a disclaimer not previously seen in reports was added to the data, and as of the article’s writing date, the death statistics for 2022 and Institutional Quality Reports, including forecasts, were still undisclosed.

Considering these limitations, four studies have been conducted in Turkey to evaluate excess deaths. Due to the lack of reliable data sharing from central public resources, all four studies relied on data provided by local governments, highlighting the importance of local data in understanding the impact of COVID-19 in the absence of centralized information. Three of these studies focus on Istanbul, the country’s most populous province, while the fourth centers on Bursa, the fourth most populous province (3–6).

In these studies, the p-score, or the percentage difference between the reported and projected number of deaths, is used to analyze excess mortality. Musellim et al. compared excess mortality in Istanbul between January 1, 2020, and May 18, 2020, to the previous five years and found a higher weekly death count with a 10% increase between the 10th and 15th weeks (March 2020 - April 2020) (3). Yardim and Eser, using municipal records from Istanbul, analyzed excess mortality from March 11, 2020, to July 12, 2020, and found a p-score of 21.3% with an excess mortality to official COVID-19 death ratio of 1.88 (4). Uçar and Arslan’s study on Istanbul examined daily all-cause mortality from the beginning of 2020 to November 11, 2021, and reported peak p-scores of 49.5% in the first wave (March 2020 - May 2020), 102.3% in the second wave (October 2020 – January 2021), and 77.6% in the third wave (February 2021 – June 2021), with an overall excess mortality to reported COVID-19 death ratio of 1.55 for Istanbul (5). Lastly, Pala et al. compared excess mortality in Bursa in 2020 to the previous five years and identified an average annual p-score of 35% (31.9% for women and 40.1% for men) (6). In Bursa, weekly death counts peaked in October-November 2020, and the majority (85.3%) of excess deaths were attributed to infectious diseases.

Excess mortality has been globally recognized as the most reliable measure to accurately assess the pandemic’s impact, as it encompasses the full consequences of the virus beyond case numbers or hospitalizations (7). It serves as a valuable measure for monitoring the virus’s impact over time and comparing its effects across different countries. The ratio between all cause deaths and reported COVID-19 deaths also facilitates regional and inter-country comparisons of underreporting.

Various factors have been explored in international studies in relation to COVID-19-related deaths and excess mortality, such as intensive care capacities (8), gender and age distributions (9–11), racial and ethnic differences (12), chronic diseases, urbanization, deprivation, and primary health care services (11). This study aims to investigate the relationship between regional and socio-economic differences and excess mortality in Turkey, examining how excess deaths from COVID-19 have changed over time and discussing factors contributing to these changes.

## Methods

In this cross-sectional ecological study, the latest death statistics data published by TURKSTAT on February 23, 2023, in Turkey are utilized to investigate the temporal and spatial distribution of excess mortality during the first two years (2020-2021) of the COVID-19 pandemic (1,2). The dataset provides monthly death data for each province, disaggregated by various causes. To estimate the projected number of deaths, we first accessed death data from the previous five years (January 2015 – December 2019), which is commonly used in the literature (13–18). A quasi-Poisson model that accounts for seasonal variations and the total population for each year was constructed to predict monthly projected deaths for 2020 and 2021. The ‘excessmort’ package in R was employed to conduct the modeling procedure (19).

To investigate the effects of regional and socio-economic differences on excess deaths, 81 Nomenclature of Territorial Units for Statistics-3 (NUTS-3) regions (provinces) were categorized based on the Socio-Economic Development Ranking of Provinces and Regions (SEGE-2017) and projected death numbers were calculated separately for each province and group (20). SEGE-2017 divides provinces into six classes, with the highest development level being 1 and the lowest being 6. SEGE, a research conducted by the Republic of Turkey Ministry of Industry and Technology using the Principal Component Analysis method, aims to reduce inter-regional socio-economic differences and includes 52 variables across various fields in its latest edition, SEGE-2017.

COVID-19 vaccination data obtained from the TURCOVID19 website, which republished the data provided by the Turkish Ministry of Health (21). The data on the number of people vaccinated at the provincial level (81 provinces) were initially published and updated hourly on the Turkish Ministry of Health’s website starting January 25, 2021. On September 13, 2021, the publication of vaccination numbers for the 81 provinces ceased, and consequently, the recording in the TURCOVID19 database was discontinued.

Excess deaths were calculated by subtracting the projected number of deaths at a specific level and period from the reported number of deaths. When reported deaths were equal to or less than the projected deaths, excess deaths were considered zero.

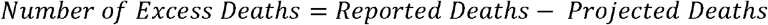

P-scores were calculated as the ratio of excess deaths to projected deaths. Calculations were performed at the national and SEGE level, both annually and monthly, and at the province (NUTS-3) level, annually.

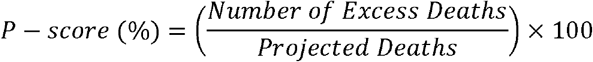

The ratio of excess death numbers to the official COVID-19 death numbers was also calculated at the national level.

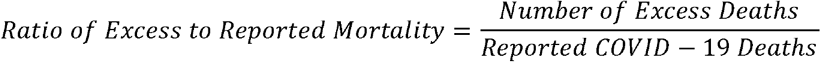

## Results

Figure 1 provides data on the number of deaths in Turkey between 2020 and 2021, along with the estimated deaths, excess deaths, and P-scores (%) by month.

**Figure 1.**
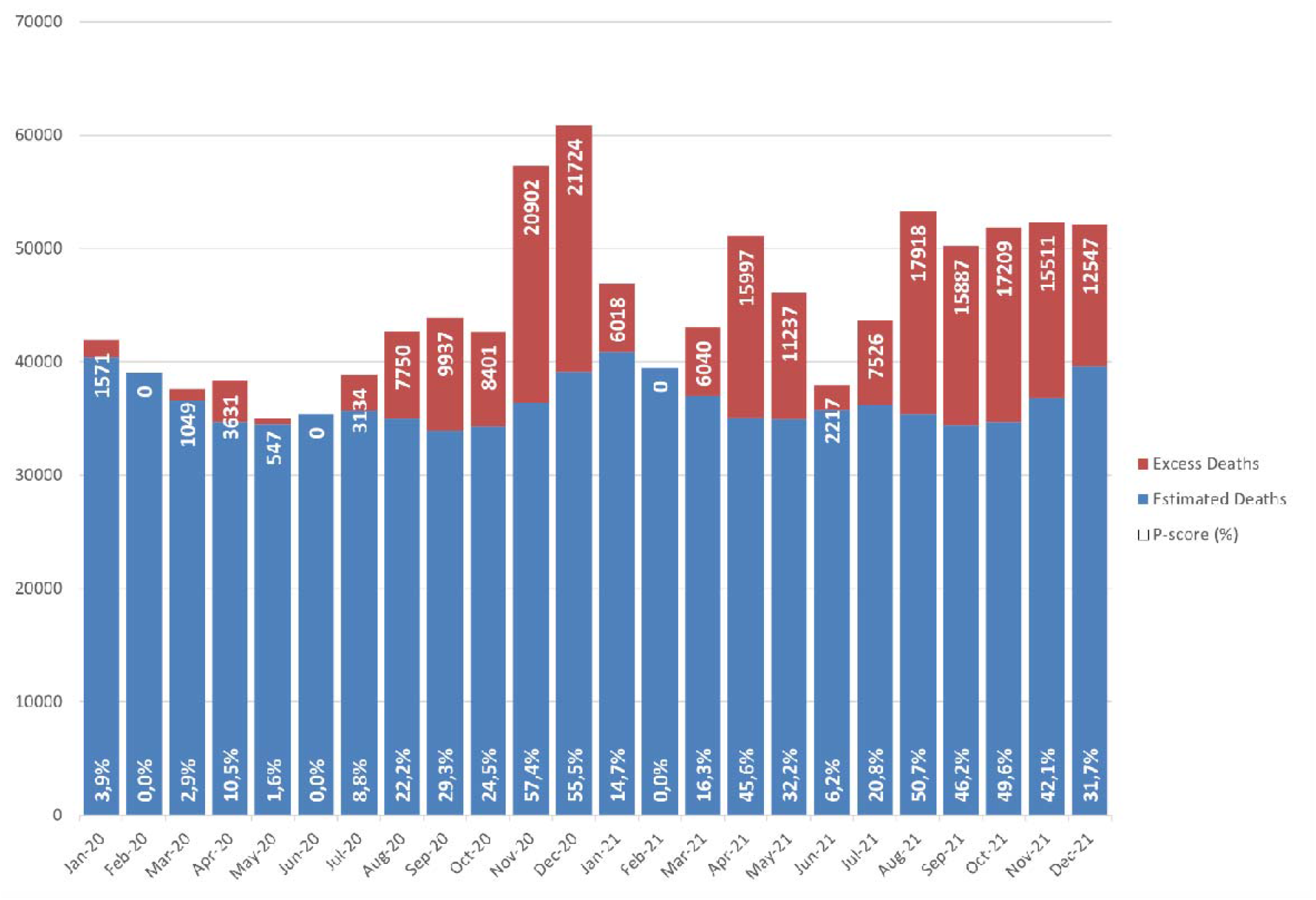
Excess Deaths due to COVID-19 in Turkey between 2020-2021 by Month

In 2020, the first cases of COVID-19 were reported in Turkey in March. From April onwards, the number of excess deaths increased significantly, with P-scores ranging from 10.5% to 57.4%. November and December were the months with the highest number of excess deaths, with P-scores of 57.4% and 55.5%, respectively.

In 2021, the excess deaths due to COVID-19 continued to be high, with rates ranging from 0% to 50.7%. August and October were the months with the highest number of excess deaths, with rates of 50.7% and 49.6%, respectively.

Figure 2 shows the cumulative excess deaths due to COVID-19 in Turkey between January 2020 and December 2021, broken down by month. Cumulative excess deaths due to COVID-19 increased steadily from March to August 2020, with August having the highest number of excess deaths. The trend continued through September and October, peaking in November. The excess deaths in December 2020 were also significant, bringing the cumulative total to 78645. The number of excess deaths increased again in March and April 2021, after having briefly decreased to zero in February.

**Figure 2.**
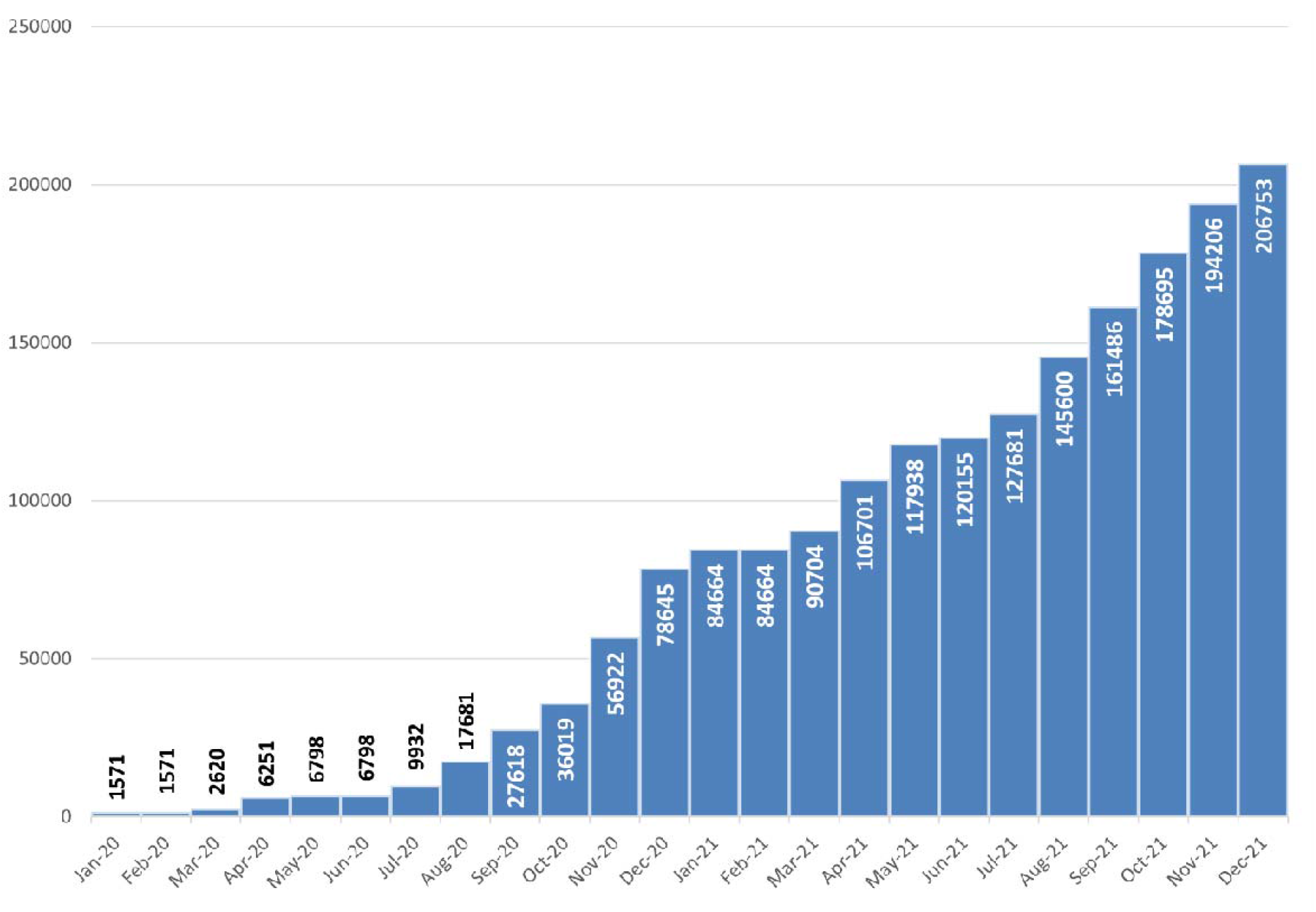
Cumulative Excess Deaths due to COVID-19 in Turkey (March 2020 - December 2021)

The highest number of excess deaths in 2020 occurred in December, with 21724 excess deaths. November 2020 also had high excess death counts. In December 2021, the excess deaths due to COVID-19 reached a cumulative total of 206753 deaths.

It is important to highlight that the officially reported COVID-19-related deaths in Turkey amounted to 22,136 in 2020 and 65,198 in 2021, totaling 87,334. The overall excess mortality to reported COVID-19 death ratio was 3.55 for 2020 and 1.96 for 2021. This finding indicates that excess deaths attributable to COVID-19 in Turkey were significantly greater than the officially reported COVID-19-related deaths during the study period.

Table 1 demonstrates the excess mortality, 95% confidence intervals (CI), and P-scores, along with their 95% CI, in Turkey during the years 2020 and 2021, stratified by Socioeconomic Indicator (SEGE) levels. The excess deaths increased from 72,886 in 2020 to 125,540 in 2021, and P-scores rose from 16.8% to 28.5%. In 2020, declared COVID-19 deaths were 22,136, while in 2021, declared COVID-19 deaths were 65,198. The data reveals a gradient in excess mortality across SEGE levels, with the highest rates observed in SEGE 6, the lowest socioeconomic group. In 2020, the excess deaths for SEGE 6 were 8,422 (25.3% P-score), while in 2021, they increased to 10,391 (31.6% P-score). The lowest rates were found in SEGE 1, the highest socioeconomic group, with 26,773 excess deaths (15.5% P-score) in 2020 and 47,452 excess deaths (26.9% P-score) in 2021.

**Table 1.** Excess Mortality and P-scores by Socioeconomic Indicator (SEGE) in Turkey during 2020-2021

| Year | Grade | Expected<br>death | Excess<br>death | Excess death<br>(%95 CI) | P-score (%) | P-score (%)<br>(%95 CI) |
| --- | --- | --- | --- | --- | --- | --- |
| 2020 | Turkey | 435052 | 72886 | (46397 - 97768) | 16.8 | (10.7 - 22.5) |
| 2021 | Turkey | 440054 | 125540 | (94303 - 154593) | 28.5 | (21.4 - 35.1) |
| 2020 | SEGE 1 | 172871 | 26773 | (18304 - 34841) | 15.5 | (10.6 - 20.2) |
| 2021 | SEGE 1 | 176237 | 47452 | (37383 - 56968) | 26.9 | (21.2 - 32.3) |
| 2020 | SEGE 2 | 81922 | 12358 | (7132 - 17264) | 15.1 | (8.7 - 21.1) |
| 2021 | SEGE 2 | 82910 | 23312 | (17135 - 29053) | 28.1 | (20.7 - 35) |
| 2020 | SEGE 3 | 64985 | 9964 | (5715 - 13949) | 15.3 | (8.8 - 21.5) |
| 2021 | SEGE 3 | 65721 | 19090 | (14071 - 23748) | 29.0 | (21.4 - 36.1) |
| 2020 | SEGE 4 | 45192 | 8963 | (5839 - 11877) | 19.8 | (12.9 - 26.3) |
| 2021 | SEGE 4 | 45307 | 13039 | (9380 - 16414) | 28.8 | (20.7 - 36.2) |
| 2020 | SEGE 5 | 36814 | 6406 | (3683 - 8931) | 17.4 | (10 - 24.3) |
| 2021 | SEGE 5 | 37037 | 12256 | (9050 - 15193) | 33.1 | (24.4 - 41) |
| 2020 | SEGE 6 | 33268 | 8422 | (5724 - 10906) | 25.3 | (17.2 - 32.8) |
| 2021 | SEGE 6 | 32842 | 10391 | (7282 - 13218) | 31.6 | (22.2 - 40.2) |

Figure 3 displays the P-scores for each month during 2020 and 2021, stratified by SEGE levels. Throughout the study period, fluctuations in P-scores can be observed among different levels. In 2020, the highest P-scores were reported in November and December, with values ranging from 45.4% to 66.7% among different SEGE levels. However, Grade 6, the lowest socioeconomic group, experienced particularly high P-scores in August (71.0%) and September (58.2%) of 2020. Similarly, in 2021, the highest P-scores were observed in April and October, with the values ranging from 20.9% to 59.6% across SEGE levels. Grade 6 exhibited notable P-scores again in August (101.7%) and September (65.9%) of 2021. Grade 1, the highest socioeconomic group, saw an early increase in excess mortality, with a P-score of 18.6% in April 2020.

**Figure 3.**
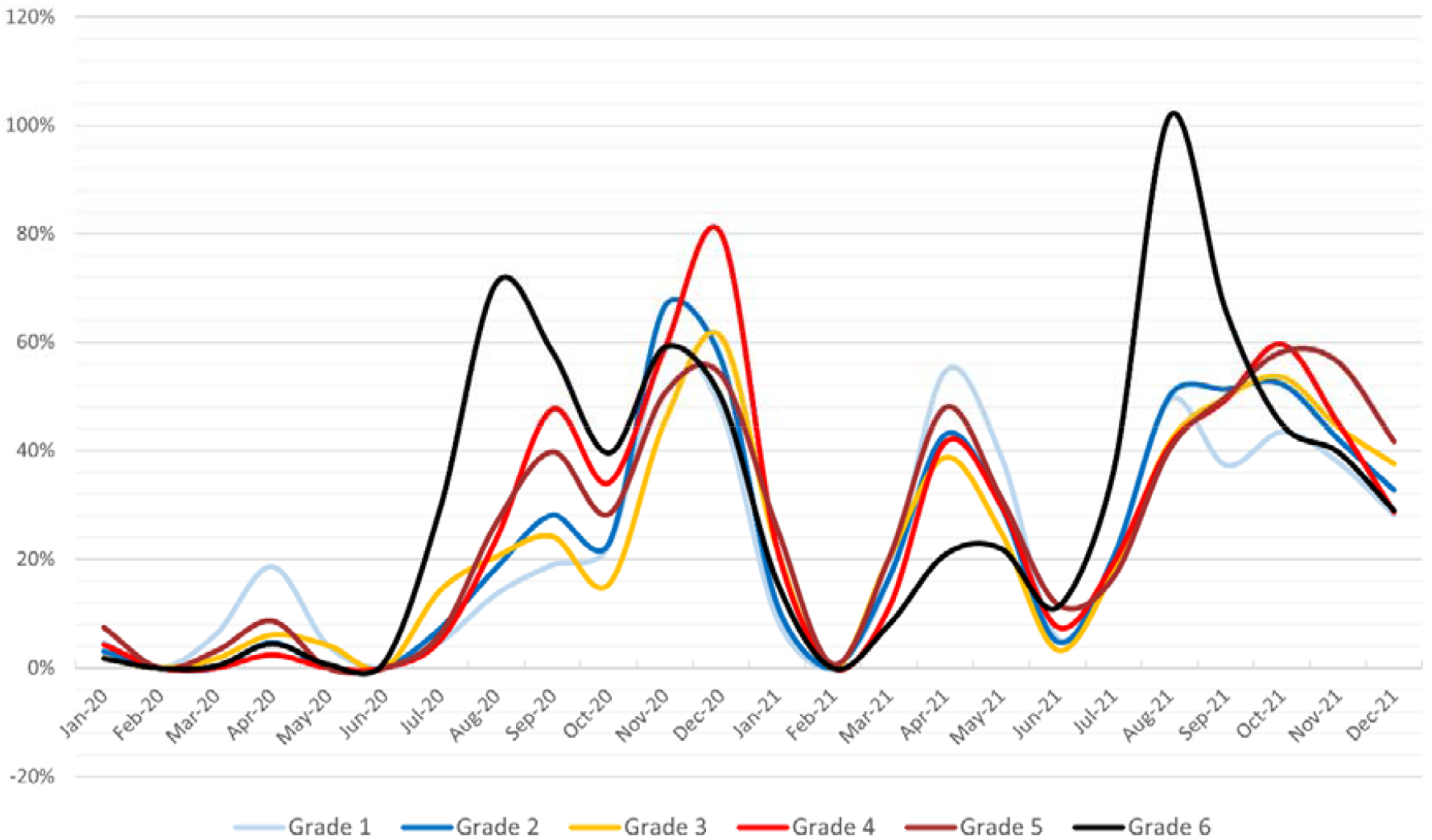
Monthly P-scores of Excess Mortality by Socioeconomic Indicator (SEGE) in Turkey during 2020-2021

Figure 4-5 is a heat map illustrating the distribution of excess mortality rates (P-scores) at the provincial level in Turkey. In 2020, when vaccination was not yet available, provinces in Southeastern Anatolia such as Şırnak, Diyarbakır, and Batman, along with Çankırı, Gümüşhane, and Bayburt, exhibited the highest excess mortality rates. In comparison, provinces on the western side of the country experienced lower excess deaths. Widespread vaccination for the elderly commenced at the beginning of 2021. The highest P-scores in 2021 were observed in provinces such as Osmaniye, Bayburt, Ağrı, Gümüşhane, Kayseri, and Amasya. Although there was an overall increase in P-scores across the country, the most notable surge occurred in the northern region (Black Sea region). Some cities, such as Uşak (19.9% to 9.9%), Hakkari (26.4% to 14.0%), and Tunceli (26.6% to 15.9%), experienced a decrease in excess mortality rates compared to the previous year.

**Figure 4.**
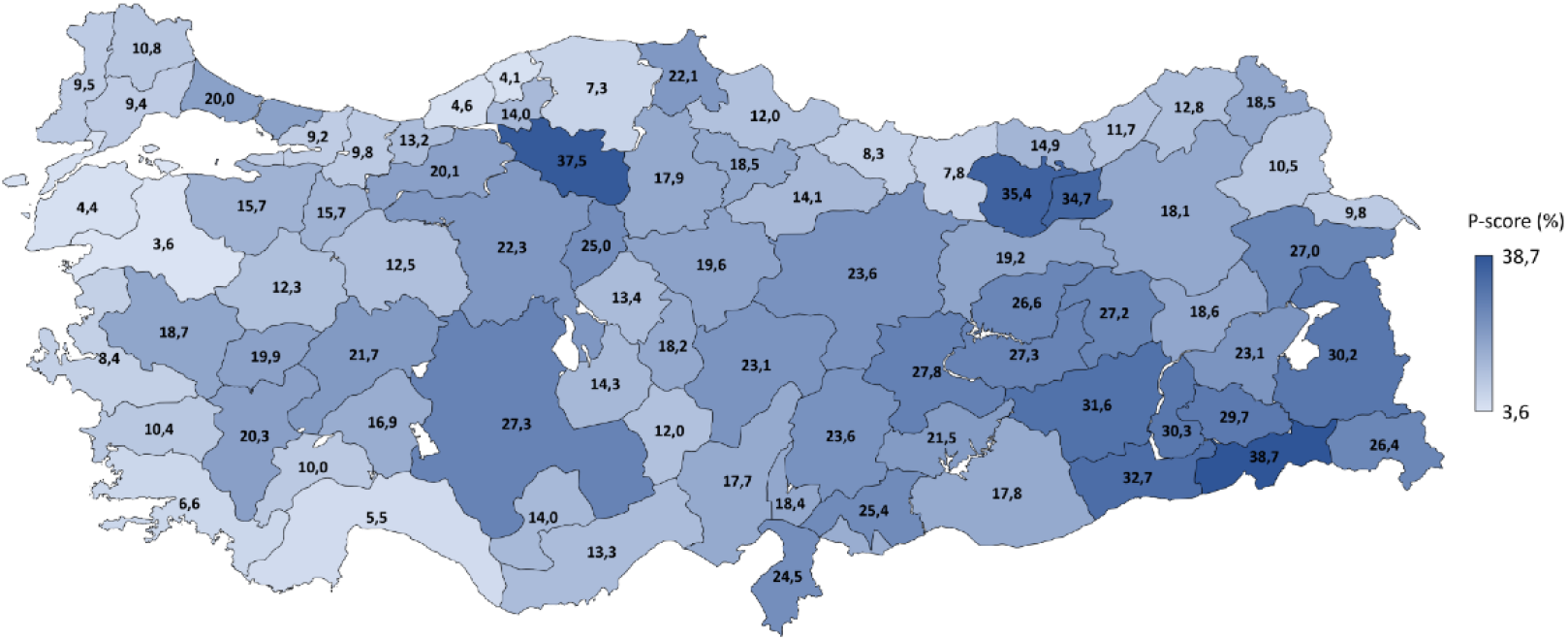
Map of Yearly P-scores by Cities in Turkey during 2020

**Figure 5.**
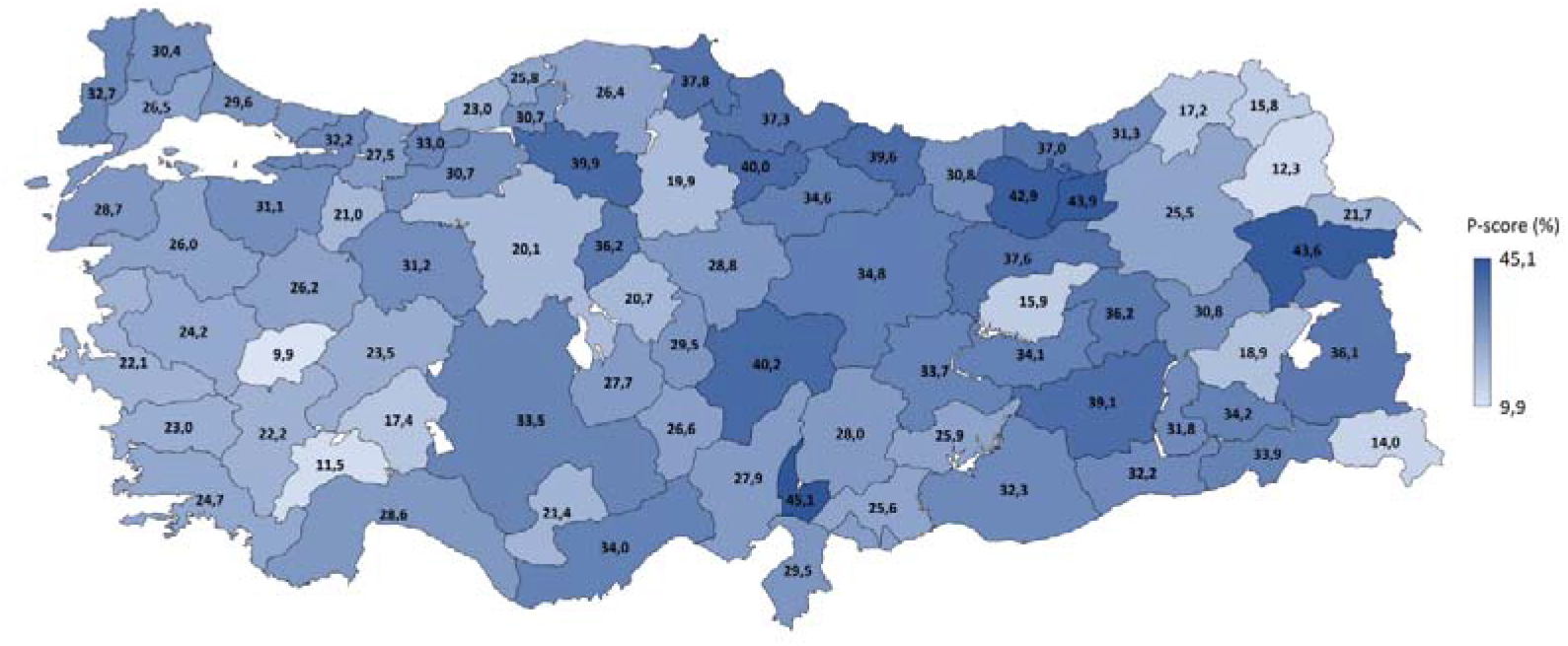
Map of Yearly P-scores by Cities in Turkey during 2021

In age distribution of all and declared COVID-19 deaths in Turkey for the years 2020 and 2021, we observed notable differences across age groups (Figure 6-7). In the year 2020, all-cause mortality reached a significant 507,938; however, a noticeable increase occurred in 2021 with the figure rising to 565,594. Notably, an age-oriented skewness characterized the dispersion of these fatalities, as older age groups - specifically, 75-84 years (142,831 in 2020, 157,896 in 2021) and 85+ years (113,539 in 2020, 125,467 in 2021) - experienced the majority of these deaths.

**Figure 6.**
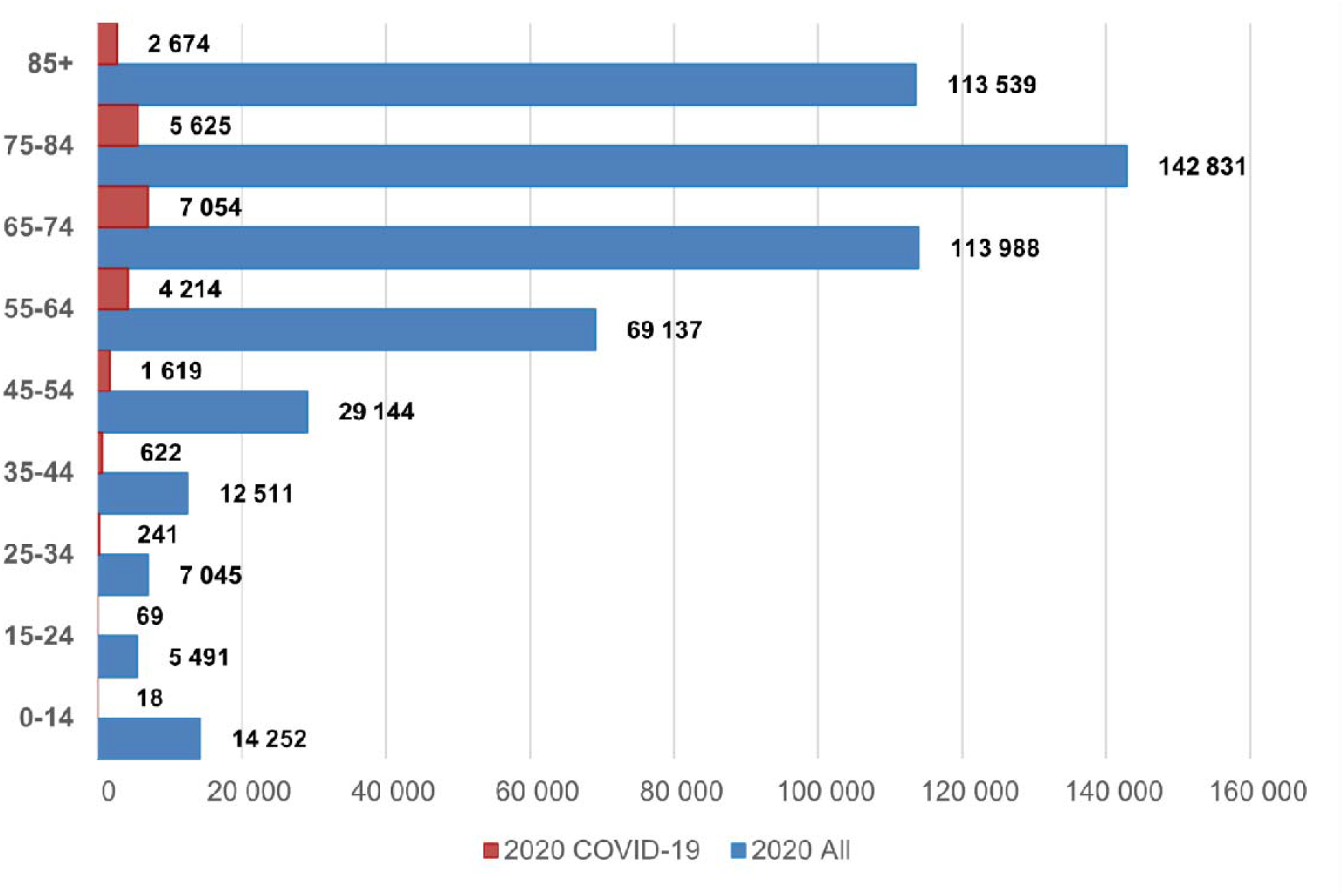
Age Distribution of All and Declared COVID-19 Deaths in Turkey (2020)

**Figure 7.**
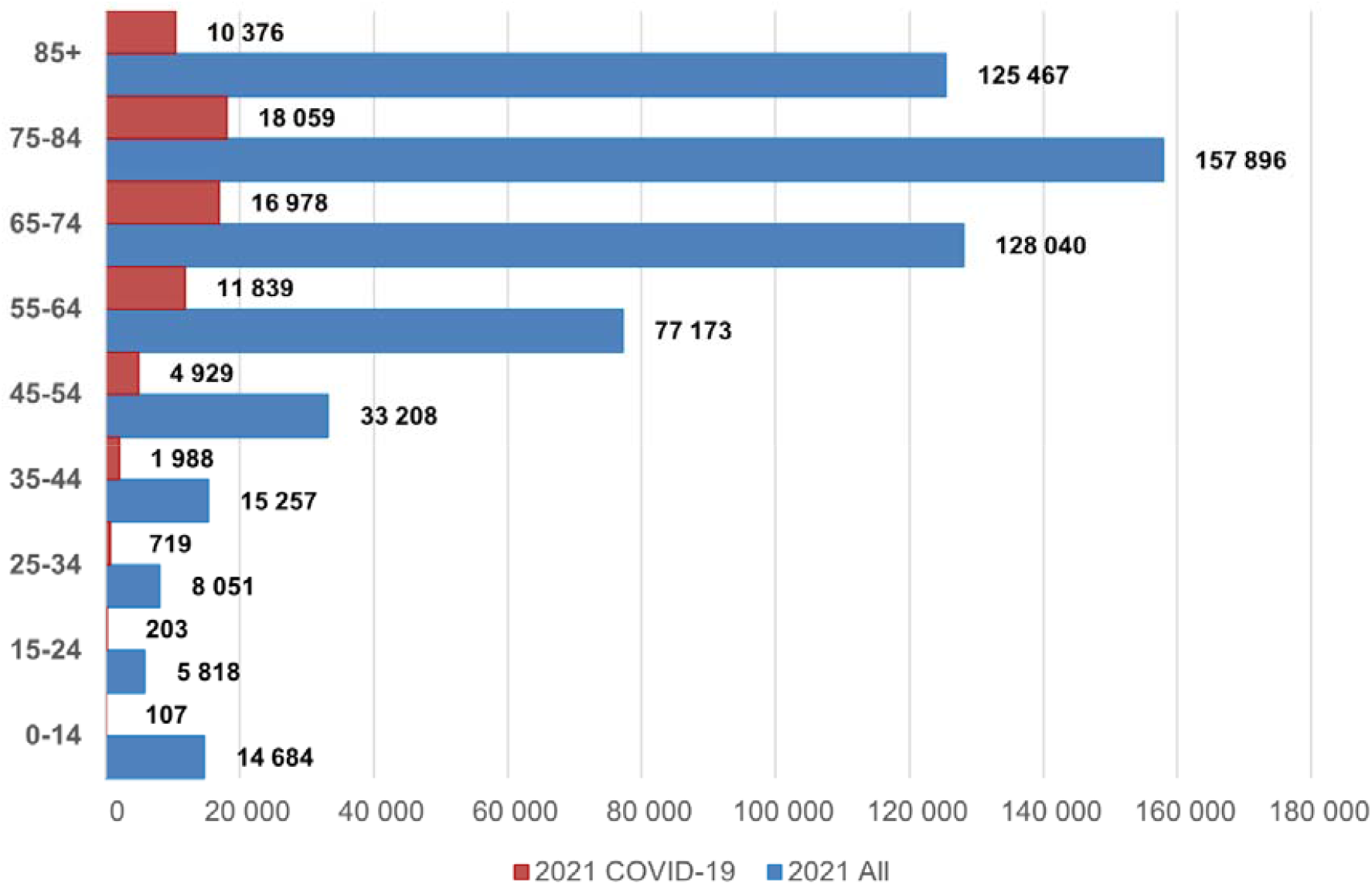
Age Distribution of All and Declared COVID-19 Deaths in Turkey (2021)

When examining declared COVID-19 deaths, a similar pattern emerged, with a higher concentration of deaths in older age groups. In 2020, there were 22,136 reported COVID-19 deaths, mainly among people aged 65-74 (7,054), 75-84 (5,625), and 85 or older (2,674). In 2021, the number of declared COVID-19 deaths increased to 65,198, again predominantly affecting the 65-74 (16,978), 75-84 (18,059), and 85+ (10,376) age categories.

It is important to note that younger age groups, while experiencing fewer COVID-19 deaths, still showed an increase in mortality rates between 2020 and 2021. In 2020, 241 COVID-19 deaths were reported in the 25-34 age group, increasing to 719 in 2021. Similarly, the 35-44 age group reported 622 COVID-19 deaths in 2020 and 1,988 in 2021. This information shows that we must think about how COVID-19 affects all age groups and be aware that the actual number of COVID-19-related deaths in younger people might be higher than reported.

It is observed that the proportion of deaths in 85+ age group was lower than all cause deaths and on the contrary proportion of COVID-19 deaths were higher in 65-74 in 2019 and 55-64 in 2020. May be due to not taking the very elderly to the hospital or that they stayed at home more than the other groups.

## Discussion

Our study provides a useful insight on the impact of COVID-19 and excess deaths in Turkey 2020 and 2021. The findings reveal a substantial rise in excess deaths during this period, attributable predominantly to pandemic. The surge in excess deaths began in April 2020, peaking in November and December with P-scores of 57.4% and 55.5%, respectively. The trend persisted into 2021, culminating in the highest number of excess deaths in August, totaling 17,918. By the end of the two-year period, the cumulative deaths amounted to 206,753. These results are consistent with previous studies in Turkey (3–6). Despite potential discrepancies between TURKSTAT data and municipal death records, the magnitude of these disparities is minimal during the pandemic.

Kontis et al. discussed the excess mortality rate between mid-February 2020 and mid-February 2021 in 40 countries (22). When compared to the overall study findings, Turkey’s excess mortality rates (16.7% in 2020, 28.5% in 2021) appear to be lower than those of some other countries. For instance, the USA, Czechia, Slovakia, and Poland experienced at least 20% higher mortality rates, while Finland, Greece, Cyprus, and Denmark experienced increases of 2-5%. In terms of excess deaths per 100,000 people, in Turkey increased from 524 in 2019 to 607 in 2020 and 667 in 2021. This indicates approximately 80 additional deaths per 100,000 people in 2020 and 143 additional deaths in 2021. Countries such as Bulgaria, Romania, Lithuania, Czechia, Poland, Slovakia, and Portugal experienced more than 200 excess deaths per 100,000 people, while Italy, the USA, England and Wales, Slovenia, Spain, Croatia, Belgium, and Montenegro experienced between 150 and 200.

It is worth noting that the overall excess mortality to reported COVID-19 death ratio in 2020 was 3.55 and 1.96 in 2021. This finding indicates that excess deaths attributable to COVID-19 in Turkey were significantly greater than the officially reported COVID-19-related deaths during the study period. Our study’s findings, which reveal that declared COVID-19 deaths accounted for only 28% of total excess deaths in 2020 and 51% in 2021 in Turkey, align with the global phenomenon of under ascertainment of mortality in infectious diseases. Whittaker et al. emphasize that under-reporting of deaths limits our understanding of the true burden of COVID-19, and that nearly 40% of deaths worldwide are unregistered (23). Mortality statistics are often incomplete and biased, which can hinder effective pandemic response and preparedness for future health crises.

The study highlights that differences within the country are better reflected by socioeconomic strata. In 2020, the highest excess deaths (25.3%) occurred in SEGE Grade 6, the lowest socioeconomic group, while in 2021, it ranked second to Grade 5 (33.1%) with 31.6%. Grade 6 largely overlaps with the Southeast region, where the heat map shows a significant excess mortality in 2020. Excess deaths in Grade 6 rose markedly during summer 2020 compared to the rest of the country and doubled the projected values in autumn 2021. Factors such as crowded households, economic challenges, limited healthcare access, and low health literacy might explain this situation. The strict lockdown for the elderly, implemented throughout 2020, may have been less effective in this region compared to the west. The region’s younger population, greater freedom of movement, and extended family structure could have facilitated the spread of the disease to older individuals. The emergence of new, highly transmissible variants and the lifting of social restrictions may have contributed to the higher excess deaths in 2021. In 2021, Turkey began widespread vaccination, prioritizing the elderly. There is a noticeable difference between the western and eastern regions in terms of total COVID-19 vaccine doses administered per 100 people. SEGE 6 has the lowest rate at 76.6 doses per 100 people, while SEGE 1 and SEGE 2 have higher rates at 126.4 and 135.5 doses per 100 people, respectively. In eastern provinces like Tunceli, which achieved high vaccination rates early in the year (151.7 doses per 100 people), excess mortality rates (15.9% in 2021) were lower compared to the region.

Notable differences across age groups were observed in the age distribution of all and declared COVID-19 deaths in Turkey for the years 2020 and 2021. An apparent increase in the overall rate of mortality was detected, and most of these fatalities was encountered by elderly cohorts. When examining declared COVID-19 deaths, a similar pattern emerged, with a higher concentration of deaths in older age groups. Despite the launch of COVID-19 vaccination campaign at the beginning of 2021, especially for elderly individuals, the total deaths in 2021 exceeded those in 2020. The restrictions imposed on citizens over the age of 65 in 2020 may explain the difference between the two years. The closure of schools during certain periods in 2020 may also have contributed to preventing the spread of the virus among younger age groups, as grandparents often take care of their grandchildren in families where both parents work, which is common in Turkey.

The study presents several limitations. Firstly, the SEGE categories are calculated based on various variables that might not be directly pertain to health and pandemics. Secondly, the absence of age standardization across provinces poses another limitation In Turkey, the average age in the eastern regions is lower than in western regions; age-adjusted statistics could potentially reveal even higher excess deaths in the east.

Excess mortality is a comprehensive measure that includes not only deaths directly caused by COVID-19 but also other factors related to the pandemic. These factors include deaths caused by the overburdening of the medical system due to the pandemic, result of limited access to healthcare resources or delayed treatment for non-COVID-19 conditions. The components of excess mortality provide a more complete picture of the pandemic’s impact on mortality rates and highlight the need for a holistic approach to addressing the pandemic’s effects. A strength of this study is the comprehensive evaluation of excess mortality across province-based and socioeconomic regions in Turkey. As COVID-19 mortality statistics were not publicly available, our analysis indirectly assesses age distribution of deaths, emphasizing the need for a holistic approach to address the pandemic’s multifaceted impact on health and well-being.

Trustworthy and reliable authorities and health organizations are crucial during public health crises such as the COVID-19 pandemic. Mistrust in these entities can result from various sources, including conflicting or misleading information, lack of transparency, and political interference. For example, during the pandemic, instances of misleading declarations of COVID-19 cases, the absence of regular reporting of statistics such as deaths and migrations, and decisions taken without relying on objective data, such as curfews announced at the last moment and transitioning schools to distance education, have contributed to the erosion of public trust in authorities and health organizations. This absence of trust could cause hesitation and indecision within the public regarding the severity of the pandemic, the security of vaccinations, and the effectiveness of public health protocols, thereby increasing the risk of infection. If the public would have known the real extent of the pandemic and the severity marked by excess deaths, it would have a positive impact on abiding the mitigation measures as well as increased uptake of vaccinations.

## Conclusion

In Turkey, an additional 206,000 deaths have been observed due to the pandemic. This situation, which is not different from general global trends, may also involve inaccuracies related to the recording of causes of deaths. Despite this, there have been some concerns about information sharing. In a public health emergency, it is crucial to develop and maintain trust with experts and the community through open communication, transparency, and collaboration.

## Data Availability

All data produced in the present study are available upon reasonable request to the authors.

## References

1. TURKSTAT. Death and Causes of Death Statistics, 2020 [Internet]. 2023 [cited 2023 Apr 9]. Available from: https://archive.is/mUCCt

2. TURKSTAT. Death and Causes of Death Statistics, 2021 [Internet]. 2023 [cited 2023 Apr 9]. Available from: https://archive.md/A2Uq6

3. Musellim B, Kul S, Ay P,Küçük FÇU, Dağlı E, Itil O, et al. Excess Mortality During COVID-19 Pandemic in İstanbul. Turk Thorac J. 2021 Mar;22(2):137–41.

4. Yardim M, Eser S. COVID-19 pandemisi ve fazladan ölümler: İstanbul örneği. Turk J Public Health. 2020 Dec 6;18(COVID-19 Special):14–24.

5. Ucar A, Arslan S. Estimation of Excess Deaths Associated With the COVID-19 Pandemic in Istanbul, Turkey. Front Public Health [Internet]. 2022 [cited 2023 Mar 28];10. Available from: https://www.frontiersin.org/articles/10.3389/fpubh.2022.888123

6. Pala K, Yürekli N, Cagac N, Turkkan A. All-cause excess mortality in 2020: The example of Bursa City in Turkey. Infect Dis Clin Microbiol. 2021 Dec 30;3:110–9.

7. Beaney T, Clarke JM, Jain V, Golestaneh AK, Lyons G, Salman D, et al. Excess mortality: the gold standard in measuring the impact of COVID-19 worldwide? J R Soc Med. 2020 Sep;113(9):329–34.

8. Oliveri RS, Gluud C, Wille-Jørgensen PA. Hospital doctors’ self-rated skills in and use of evidence-based medicine - a questionnaire survey. J Eval Clin Pract. 2004 May;10(2):219–26.

9. Signorelli C, Odone A, Gianfredi V, Bossi E, Bucci D, Oradini-Alacreu A, et al. COVID-19 mortality rate in nine high-income metropolitan regions. Acta Bio Medica Atenei Parm. 2020;91(9-S):7–18.

10. Kontis V, Bennett JE, Rashid T, Parks RM, Pearson-Stuttard J, Guillot M, et al. Magnitude, demographics and dynamics of the effect of the first wave of the COVID-19 pandemic on all-cause mortality in 21 industrialized countries. Nat Med. 2020 Dec;26(12):1919–28.

11. Pilkington H, Feuillet T, Rican S, de Bouillé JG, Bouchaud O, Cailhol J, et al. Spatial determinants of excess all-cause mortality during the first wave of the COVID-19 epidemic in France. BMC Public Health. 2021 Nov 24;21(1):2157.

12. Xian Z, Saxena A, Javed Z, Jordan JE, Alkarawi S, Khan SU, et al. COVID-19-related state-wise racial and ethnic disparities across the USA: an observational study based on publicly available data from The COVID Tracking Project. BMJ Open. 2021 Jun 1;11(6):e048006.

13. Alicandro G, Remuzzi G, Vecchia CL. Italy’s first wave of the COVID-19 pandemic has ended: no excess mortality in May, 2020. The Lancet. 2020 Sep 12;396(10253):e27–8.

14. Michelozzi P, de’Donato F, Scortichini M, Pezzotti P, Stafoggia M, De Sario M, et al. Temporal dynamics in total excess mortality and COVID-19 deaths in Italian cities. BMC Public Health. 2020 Aug 27;20(1):1238.

15. Karlinsky A, Kobak D. Tracking excess mortality across countries during the COVID-19 pandemic with the World Mortality Dataset. eLife [Internet]. 2021 Jun 30;10. Available from: http://dx.doi.org/10.7554/eLife.69336

16. Morgan D, Ino J, Paolantonio GD, Murtin F. Excess mortality: Measuring the direct and indirect impact of COVID-19 [Internet]. Paris: OECD; 2020 Oct [cited 2023 Apr 9]. Available from: https://www.oecd-ilibrary.org/social-issues-migration-health/excess-mortality_c5dc0c50-en

17. Weinberger DM, Chen J, Cohen T, Crawford FW, Mostashari F, Olson D, et al. Estimation of Excess Deaths Associated With the COVID-19 Pandemic in the United States, March to May 2020. JAMA Intern Med. 2020 Oct 1;180(10):1336.

18. Scortichini M, Schneider dos Santos R, De’ Donato F, De Sario M, Michelozzi P, Davoli M, et al. Excess mortality during the COVID-19 outbreak in Italy: a two-stage interrupted time-series analysis. Int J Epidemiol. 2020 Dec 1;49(6):1909–17.

19. Acosta RJ, Irizarry RA. A Flexible Statistical Framework for Estimating Excess Mortality. Epidemiology. 2022 May;33(3):346.

20. Acar S, Bilen Kazancık L, Meydan MC, Işık M. Sosyo-Ekonomik Gelişmişlik Sıralaması Araştırmaları (SEGE) 2017 [Internet]. Ankara: T.C. Sanayi ve Teknoloji Bakanlığı; 2019 [cited 2023 Apr 9]. Available from: https://www.sanayi.gov.tr/merkez-birimi/b94224510b7b/sege

21. Uçar A, Arslan Ş, Manap H, Gürkan T, Çalişkan M, Dayioğlu A, et al. Türkiye’de Covid-19 Pandemisinin Monitörizasyonu Için Interaktif Ve Gerçek Zamanlı Bir Web Uygulaması: TURCOVID19. Anatol Clin J Med Sci. 2020 Mar 20;25(Special Issue on COVID 19):154–5.

22. Kontis V, Bennett JE, Parks RM, Rashid T, Pearson-Stuttard J, Asaria P, et al. Lessons learned and lessons missed: impact of the coronavirus disease 2019 (COVID-19) pandemic on all-cause mortality in 40 industrialised countries and US states prior to mass vaccination. Wellcome Open Res. 2022 Feb 15;6:279.

23. Whittaker A, Anson M, Harky A. Neurological Manifestations of COVID-19: A systematic review and current update. Acta Neurol Scand. 2020 Jul;142(1):14–22.

